# No SARS-CoV-2 carriage observed in children attending daycare centers during the first weeks of the epidemic in Belgium

**DOI:** 10.1101/2020.05.13.20095190

**Authors:** Stefanie Desmet, Esra Ekinci, Ine Wouters, Bram Decru, Kurt Beuselinck, Surbhi Malhotra-Kumar, Heidi Theeten

## Abstract

To gain knowledge about the role of young children attending daycare in the SARS-CoV-2 epidemic, a random sample of children (n=84) aged between 6 and 30 months attending daycare in Belgium was studied shortly after the start of the epidemic (February 29^th^) and before the lockdown (March 18^th^). No asymptomatic carriage of SARS-CoV-2 was detected, whereas common cold symptoms were common (51.2%).

## Main text

Understanding the burden of carriage in children and the subsequent potential for transmission of severe acute respiratory syndrome-associated coronavirus (SARS-CoV-2) is important to implement public health measures and to decide on the exit strategy for the current lockdown in Belgium. This study investigated the (asymptomatic) carriage of SARS-CoV-2 in children attending daycare centers during the period 2-12 March 2020 to get more insight into the possible role of children in the transmission of SARS-CoV-2 during the first weeks of the SARS-CoV-2 epidemic in Belgium. Although the first confirmed case was on February 4^th^ 2020, the real start of the epidemic was detected from February 29^th^ onwards with a total of 689 confirmed cases spread over the whole country on March 13^th^ [1].

## Why we studied SARS-CoV-2 carriage in young children

In December 2019, SARS-CoV-2 has emerged in Wuhan, China, causing the 2019 novel coronavirus disease (COVID-19). This virus eventually spread worldwide and in March 2020 the World Health Organization (WHO) announced SARS-CoV-2 a pandemic health emergency. Person-to-person transmission of SARS-CoV-2 takes place mainly through close contact with an infected person (mainly via respiratory droplets) and after touching infected objects [2]. Clinical manifestations of SARS-CoV-2 are fever, dry cough, shortness of breath and respiratory failure and vary from very mild to severe symptoms [3].

Current evidence indicates that older populations are most susceptible to severe presentations of COVID-19, however, there is less knowledge about disease severity and transmission amongst infants and children [4]. Evidence in China suggests that the clinical symptoms of COVID-19 may be less severe in children and thus harder to recognize [5, 6]. Possible explanations could be: 1) children might have fewer opportunities for exposure to pathogens or infected patients [5], 2) lower expression of the Angiotensin converting enzyme II (ACE 2) receptor as it is indicated that ACE 2 is likely the receptor for SARS-CoV-2 [7], 3) children often get respiratory infections (including Coronaviruses) in the winter and might therefore have higher antibody levels than adults [5], 4) the immune system of children is still developing and may react differently on pathogens compared to adults' immune system [8].The lower symptomatic disease incidence in the pediatric population raises the concern that this population could be an important source of SARS-CoV-2 transmission [6].

### Ethical statement

The current study was in line with the Declaration of Helsinki, as revised in 2013. Approval to conduct the current study with ID 18/31/355 was obtained from the University of Antwerp and University Hospital of Antwerp ethics committee (Commissie voor Medische Ethiek van UZA/UA) on 29/07/2019. The informed consent allowed us to determine other respiratory pathogens.

## Study population and sampling

The current study was embedded in the nasopharyngeal carriage study that started in Belgium in 2016 to monitor changes in the proportions of pneumococcal serotypes in children between six and thirty months of age, attending daycare centers (DCCs) [9]. DCCs were randomly selected throughout Belgium. After written consent of at least one parent, a single nasopharyngeal (NP) swab was collected. A questionnaire regarding the child's demographic and clinical characteristics, as well as pneumococcal vaccination status was filled in by their parents. Signs of common cold in children were defined as coughing and/or running nose, and were registered at the moment of sampling [9].

Sample collection during 2019-2020 was performed from beginning November 2019 to the end of March 2020. This collection period spanned the crucial first weeks of the COVID-19 epidemic in Belgium, thus enabling us to study the introduction, if any, of the SARS-CoV-2 virus in the daycare population. To determine the SARS-CoV-2 carriage, NP swabs taken from 84 children attending 8 different daycare centers (1 in Brussels, 3 in Wallonia and 4 in Flanders) during the period of 2-12 March 2020 were analyzed. Of the 84 children included in this study, relevant population characteristics were: 1) 43 (52.4%) were girls; 2) at the moment of sampling, signs of common cold were observed in half of the children (51.2%); 3) just over half (56.1%) of the children had at least one sibling in the same household; 4) the majority of children (87.8%) stayed at least twice a week in daycare; and 5) the parents of the majority of the children (75.6%) were non-smokers.

## In-house SARS-CoV-2 real-time PCR

The collected swabs were transported on dry ice and stored in 1mL STGG (Skim milk – Tryptone – Glucose – Glycerol). In-house SARS-CoV-2 real-time PCR was performed on 200 μL of the sample according to WHO guidelines [10]. Validation of the test on STGG medium was performed by adding 3μL of COVID-19 positive sample (n=3) to 300μL STGG media, followed by storage in a refrigerator (4°C) for 24 hours and afterwards freezing at -80°C. Ct-values of the frozen STGG samples were similar as the Ct-values of initial sample (1/100 diluted).

## Results do not suggest a role of daycare attendance in early transmission

All analyzed samples were negative for SARS-CoV-2, which means that shortly after the start of the epidemic (February 29^th^) and before the lockdown in Belgium (March 18^th^) no (asymptomatic) carriage of SARS-CoV-2 was detected in a random sample of children (n=84) aged between 6 and 30 months attending daycare. Only one sample had an amplification curve (Ct value of 38.8). To confirm this weak signal, extract of this sample was reanalyzed in triple and no amplification was observed. A limitation of our study is that we have no information on COVID-19-like symptoms in household members or caregivers.

The result is in line though with other studies that suggest only a minimal role of children in the epidemic of SARS-CoV-2. Iceland reported children under 10 years of age less likely to be positive by RT-PCR testing than were persons 10 years of age or older in large scale screening in a random sample of the general population (n=13080 of whom 848 children were all negative vs. 0.8% positives in the remaining sample) [11]. Similarly, in Italy no children younger than 10 were infected in a large-scale survey before and after the start of the lock-down in the municipality where the first COVID-19 death in Italy was reported [12]. Among confirmed COVID-19 patients, the percentage of children was as low as 1-5% in studies published up to March 18 2020, and in United states up to April 10, [13, 14]. Although children in China had a similar risk of infection as adults [15], China reported a lower secondary attack rate to children than to adults (4% vs 17%) in a household study [16]. Only a handful of deaths among children with COVID-19 have been reported worldwide and generally symptoms in children are mild [17]

Limited evidence from contact research suggests that children less frequently infect other people than do adults, although similar viral loads as in adults were observed in an unpublished German study [18]. In the Netherlands, a household study showed that the spread of SARS-CoV-2 occurs mainly between people of approximately the same age, and that patients under 20 years of age had a smaller impact on the spread of the virus compared to adults [19]. A recent review found only 9.7% pediatric index cases among 31 intra-household transmission clusters identified from literature [20]. Our study adds that in Belgium, where the epidemic was imported mainly by adult travelers, there is no sign of early introduction into daycare centers at a moment children were not yet isolated at home although the virus was clearly circulating. It is clear that more evidence is needed to understand the actual role of young children in transmission of SARS-CoV-2 and their infection risk when attending daycare.

## Data Availability

The data that support the findings of this study are available from the corresponding author,[HT].

